# Prevalence of Depressive symptoms and its associated factors among elderly living in old age homes of Kathmandu Metropolitan City, Nepal

**DOI:** 10.1101/2024.05.29.24308142

**Authors:** Sanju Maharjan, Nujan Tiwari, Sita Bista, Prem Basel

## Abstract

**Background:** Depressive symptoms have become a global public health problem, with a predominant effect on the elderly people. The studies on mental health status of elderly people in Nepal are quite limited. In this study, we aim to assess the prevalence of depressive symptoms and its associated factors among the elderly living in public old age homes of Kathmandu Metropolitan City in Nepal.

**Methodology:** A cross-sectional study was conducted including 142 adults aged 60 years and above recruited through proportional simple random sampling from six old age homes. The 15 item-Geriatric Depression Scale(GDS) was used to assess the depressive symptoms among the participants. Associated factors were tested using Chi-square test; and a p-value of less than 0.05 with a confidence interval of 95% was used for statistical significance.

**Results:** More than half of the study participants, 58.5% (95% CI: 49.9%-66.7%) were found to have depressive symptoms. Among them, 38.7% had mild symptoms, 16.2% had moderate symptoms and 3.5% had severe symptoms. Age (OR = 2.25, 95% CI: 1.08-4.66), sex (OR = 2.36, 95% CI: 1.17-4.75), past family type (OR = 0.44, 0.22-0.89), chronic physical health problem (OR = 0.34, 95% CI: 0.12-0.98) and feelings of loneliness were found to have significant association with depressive symptoms among the elderly population.

**Conclusion:** The prevalence of depressive symptoms among the elderly in old age homes in Kathmandu Metropolitan City is quite high and a concerning issue that requires targeted mental health programs and interventions in order to bring about a positive shift in their mental health condition.

## Introduction

The World Health Organization (WHO) defines depression as a mental health disorder that presents with persistently depressed mood, loss of interest or pleasure in previously pleasurable activities, decreased energy, feelings of guilt or low self-worth, disturbed sleep or appetite, and poor concentration. (1). Depressive symptoms can affect the quality of life and is one of the leading causes of disability worldwide(2,3) Everyone is susceptible to mental health problems, whatever stage of life they might be at. However, the elderly are at an even higher risk of mental health problems including anxiety and depression, with depression being the most common psychiatric disorder in later life(3,4)

Deep rooted social stigma, misconceptions and serious inadequacies with regards to information, resources and health facilities, all act as barriers to accessing proper mental health services, especially in developing countries. Nepal government has limited number of trained human resources and funds for the effective and efficient implementation of the National policy, act and regulations on ageing and the problems of the elderly. As such, the unique health needs of elderly people have largely been neglected(5). Further, a 3.5% population growth rate of the elderly is a challenge to the existing healthcare demand in the country. (6)

While it is true that additional years of life open up a wide range of opportunities and contributions to society, we cannot be oblivious to the fact that it all depends on the health of the population. (7). Multiple risk factors leading to mental health problems at any point in life have been identified but the stressors that have been deemed significant among elderly people include depreciation of physical, social and cognitive functions from illness, retirement, bereavement, loss of income and disability(7,8). The presence of one or more of these risk factors often result in isolation, loneliness and distress in elderly people that require a significant amount of care(3,7). They are commonly perceived as a burden to family and society, making them all the more vulnerable to physical and emotional abuse, abandonment and financial crisis. This compels them to opt for other places to live a free and respectful life, and old age homes are often their only support to do so(4,9). Depressive symptoms have been found to be more common in institutionalized old people. As these symptoms usually coexist with other organic health problems, mental health symptoms are overlooked and often left untreated(3,8).

There have been a few studies conducted in Nepal among the elderly. In a study conducted by Chalise, the prevalence of depression among elderly living in old age homes of Devghat area of Nepal was found to be 57.8%.(10). A study by Ghimire et al comparing the prevalence of depression between elderly living in old age homes and those living in the community reported 52.73% prevalence in the old age homes which was twice of that in the community; 25.45%(11). Another study by Dhungana et al found presence of depression among 80.7% of the elderly participating in their study(12) These studies suggested that age, gender, ethnicity, poor perception of life, poor social relationships, chronic organic diseases, lack of entertainment activities were associated with higher levels of depression. Kathmandu Metropolitan City houses the largest and most diverse population demographics in the country and also has the greatest number of operational old age homes in Nepal. There is limited data regarding the prevalence of depression in these institutions. Thus, this study aims to assess the prevalence of depressive symptoms and its associated factors among the elderly people living at old age homes in Kathmandu Metropolitan City, Nepal.

## Methodology

### Study design and participants

This is an institutional based cross-sectional study conducted in six old age homes in Kathmandu Metropolitan City. The conceptualization and literature of the study was conducted from December 2021. After receiving ethical approval from the Institutional Review Board (IRB) of IOM on 16 March 2022 [Approval number: 371 (6–11) E^2^ 078/079], participant recruitment and data collection was conducted from 22 March 2022 to 5 April 2022. The study population consists of the adults of age 60 years and over, of all public old age homes in the study site. The elderly who could not communicate verbally and who did not understand Nepali language were excluded from the study.

### Sampling procedure

There are 6 old age homes working in coordination with Kathmandu Metropolitan City. According to the Metropolitan office, the total number of elderly living there is 198 and the total sample size of this study is 142. Proportionate simple random sampling was applied to select the participants. The proportion of sample from the total population is 71.72%. There are 22, 9, 7, 31, 31, 98 elderly living in 6 old age homes of Kathmandu Metropolitan City. So, the sample proportion for the study in each old age home was 16, 7, 5, 22, 22, and 70 respectively. The participants were selected randomly.

### Study variables

#### Dependent variable

Depression

#### Independent variables

Socio-demographic characters of the elderly: Age, sex, ethnicity, marital status, education, past family type, present source of income Health status: Presence of chronic medical illnesses Individual factors: Feelings of loneliness.

### Study tools

Data collection was done with face-to-face interview with selected participants from the representative sample. The tool was pretested in one of the old age homes outside the study area. Structured and validated short form of Geriatric Depression Scale (GDS 15) was used to assess depressive symptoms.(13)It consists of a 15 ‘Yes/No’ questions to measure depressive symptoms. Scoring range of 0 to 4 is considered to be normal, 5 to 8 is mild, 9 to 11 is moderate and 12 to 15 indicates severe depression.

### Data management and analysis

Collected data was entered in Epidata version 3.1 software and exported to SPSS v20 for further analysis. The questions were scored accordingly to calculate the total score and level of depressive symptoms. For further analysis, the variables were dichotomized into normal and presence of depressive symptoms. The association between depressive symptoms and different variables was tested using the chi-square test. If p-value was less than 0.05 at a 95% confidence interval, then the association between dependent and independent variables was considered statistically significant.

### Ethical consideration

Ethical approval was obtained from the Institutional Review Committee (IRC) of the Institute of Medicine (IOM). Official letter of cooperation from the Central Department of Public Health was written to the administrative office of the Kathmandu Metropolitan City and permission was also obtained from each of the old age homes regarding data collection from their residents. Verbal informed consent was obtained from all study participants to allow the use of anonymous data in research. Confidentiality of the information was maintained.

## Results

Table 1 summarizes the socio-demographic characteristics of the respondents. A total of 142 elderly participated in the study. The mean age of the participants was 76.37 years (SD: 8.21 years). The minimum age of the respondents was 60 years while the maximum age was 100 years. Majority of the participants of our study were females (64.1%), with 35.9% being males. 52.8% of the participants were of Brahmin/Chhetri ethnicity while the rest were Janajatis (indigenous groups). A majority of the elderly (61.3%) had already lost their partners during the study. More than half, 56.3% lived with joint family in the past and 57% had at least one source of income. 90.1% of the participants could not read and write.

**Table 1.** Socio-demographic characteristics of the respondents n=142.

| Characteristics | Number | Percentage |
| --- | --- | --- |
| **Age in years (Mean (± SD) = 76.37±8.21)** |  |  |
| 60-79 | 91 | 64.1 |
| 80 and above | 51 | 35.9 |
| **Sex** |  |  |
| Male | 51 | 35.9 |
| Female | 91 | 64.1 |
| **Ethnicity** |  |  |
| Janajati | 67 | 47.2 |
| Brahmin/Chhetri | 75 | 52.8 |
| **Marital status** |  |  |
| Single | 22 | 15.5 |
| Married | 17 | 12.0 |
| Widow/widower | 87 | 61.3 |
| Separated/divorced | 16 | 11.3 |
| **Education status** |  |  |
| Can read and write | 14 | 9.9 |
| Cannot read and write | 128 | 90.1 |
| **Past family type** |  |  |
| Nuclear | 62 | 43.7 |
| Joint | 80 | 56.3 |
| **Source of income** |  |  |
| Yes | 81 | 57.0 |
| No | 61 | 43.0 |
| **Type of source\*** |  |  |
| Pension | 1 | 1.1 |
| Old age allowance | 72 | 79.1 |
| Savings | 3 | 3.3 |
| Business | 15 | 16.5 |

\*Multiple response

With regards to their physical health; the majority, 63.2% had musculoskeletal problems, followed by hypertension (26.4%), gastro-enteric problems (21.6%), diabetes mellitus (21.6%), respiratory problems (20.8%) and others (4%).

### Health status of the respondents

Table 2 shows that a large group, 88% of the respondents had chronic health problems.

**Table 2.** Health status of the respondents n=142.

| **Characteristics** | **Number** | **Percentage** |
| --- | --- | --- |
| **Chronic health problems** |  |  |
| Yes | 125 | 88 |
| No | 17 | 12 |
| **Type of Problem\*** |  |  |
| Gastroenteric problems | 27 | 21.6 |
| Respiratory problems | 26 | 20.8 |
| Hypertension | 33 | 26.4 |
| Diabetes | 27 | 21.6 |
| Musculoskeletal problems | 79 | 63.2 |
| Others | 5 | 4 |

*\*Multiple response*

**Table 3.** Individual factor of the respondents n=142.

| Characteristics | Number | Percentage |
| --- | --- | --- |
| **Feeling of loneliness** |  |  |
| Yes | 65 | 45.8 |
| No | 77 | 54.2 |

### Individual factor of the respondents

Feeling of loneliness was studied under individual factor. 45.5% of the respondents shared that they felt lonely.

### Prevalence of depressive symptoms among elderly living in old age homes

The study revealed overall prevalence was found to be 58.5% (95% CI: 49.9%-66.7%). 38.7% elderly were found to have mild depressive symptoms, followed by 16.2% with moderate symptoms and 3.5% with severe depressive symptoms.

### Association of risk factors with depressive symptoms

Table 5 represents the association of depressive symptoms with different variables. It was found that age, sex and past family type were statistically significant with depressive symptoms. Elderly of age 80 and above were 2.25 times more likely to have depressive symptoms than those of age 60-79 (OR = 2.25, 95% CI: 1.08-4.66). Similarly, females were 2.36 times more likely to have symptoms than males (OR = 2.36, 95%CI: 1.17-4.75). Likewise, elderly who previously lived with joint families were found to be protected against depressive symptoms in comparison to those living in nuclear families (OR = 0.44, 0.22-0.89).

**Table 4.** Prevalence of depressive symptoms among elderly n=142.

| Level of depressive symptoms | Number | Percentage |
| --- | --- | --- |
| Normal | 59 | 41.5 |
| Mild | 55 | 38.7 |
| Moderate | 23 | 16.2 |
| Severe | 5 | 3.5 |

**Table 5.**
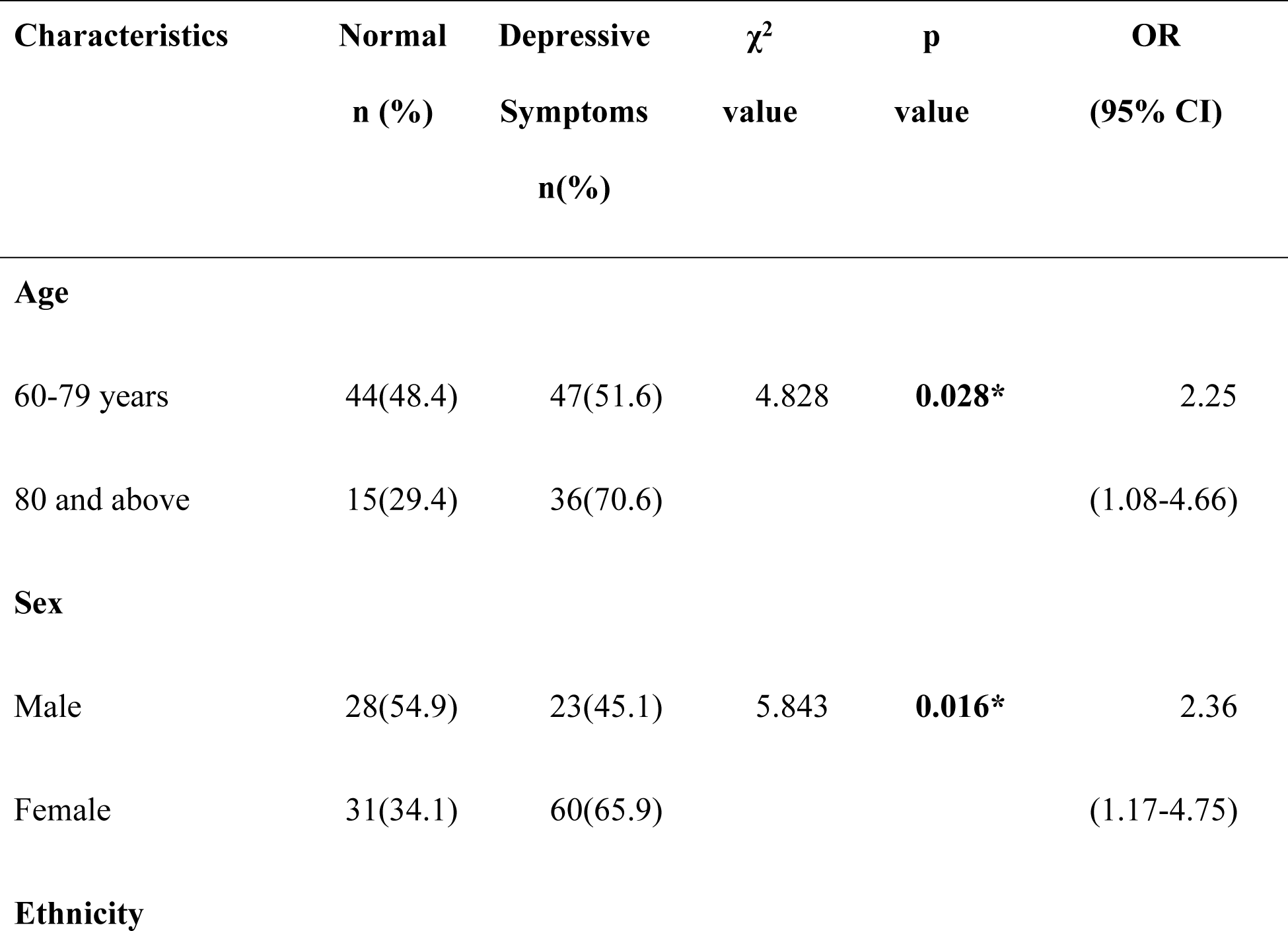
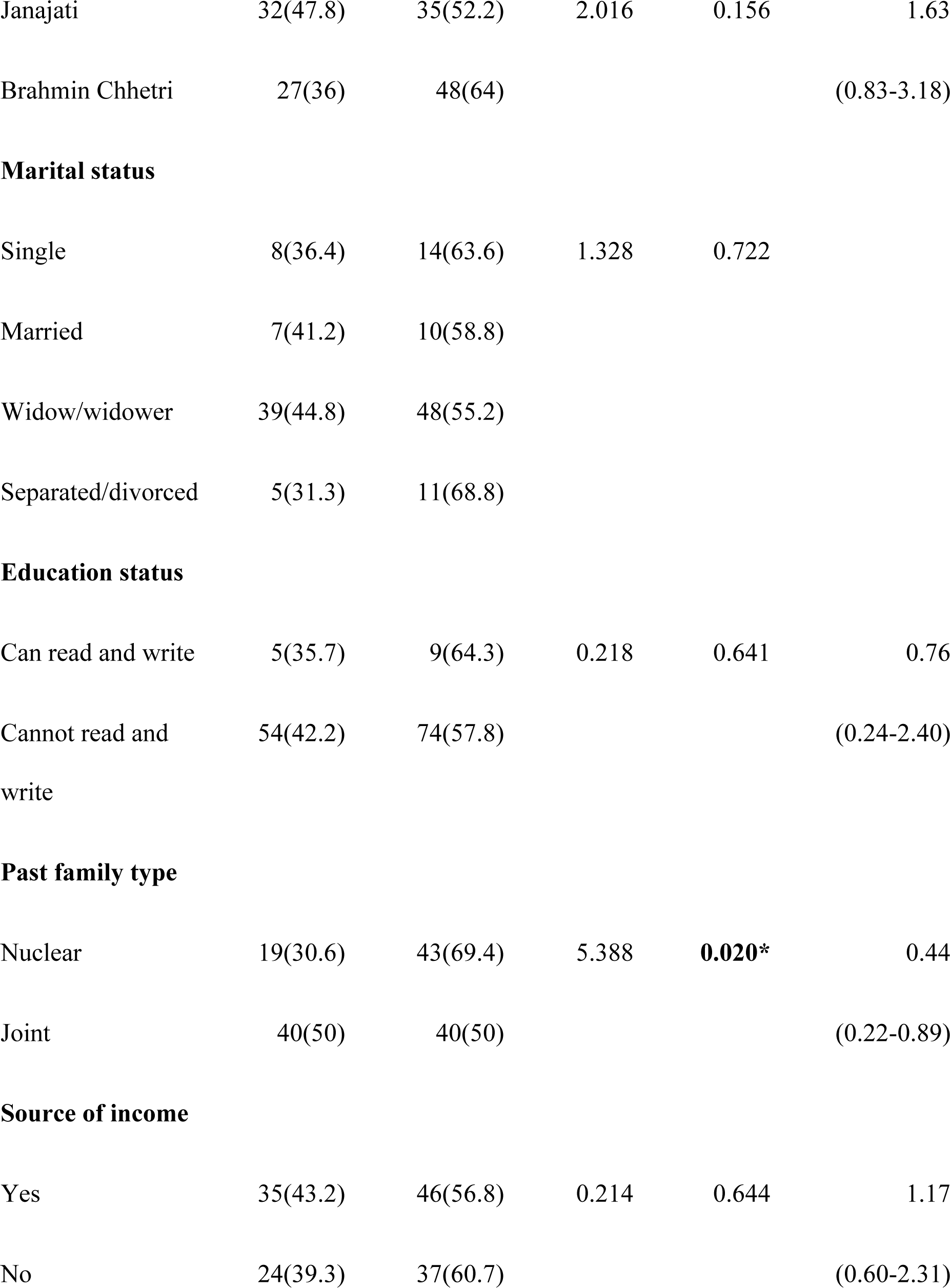
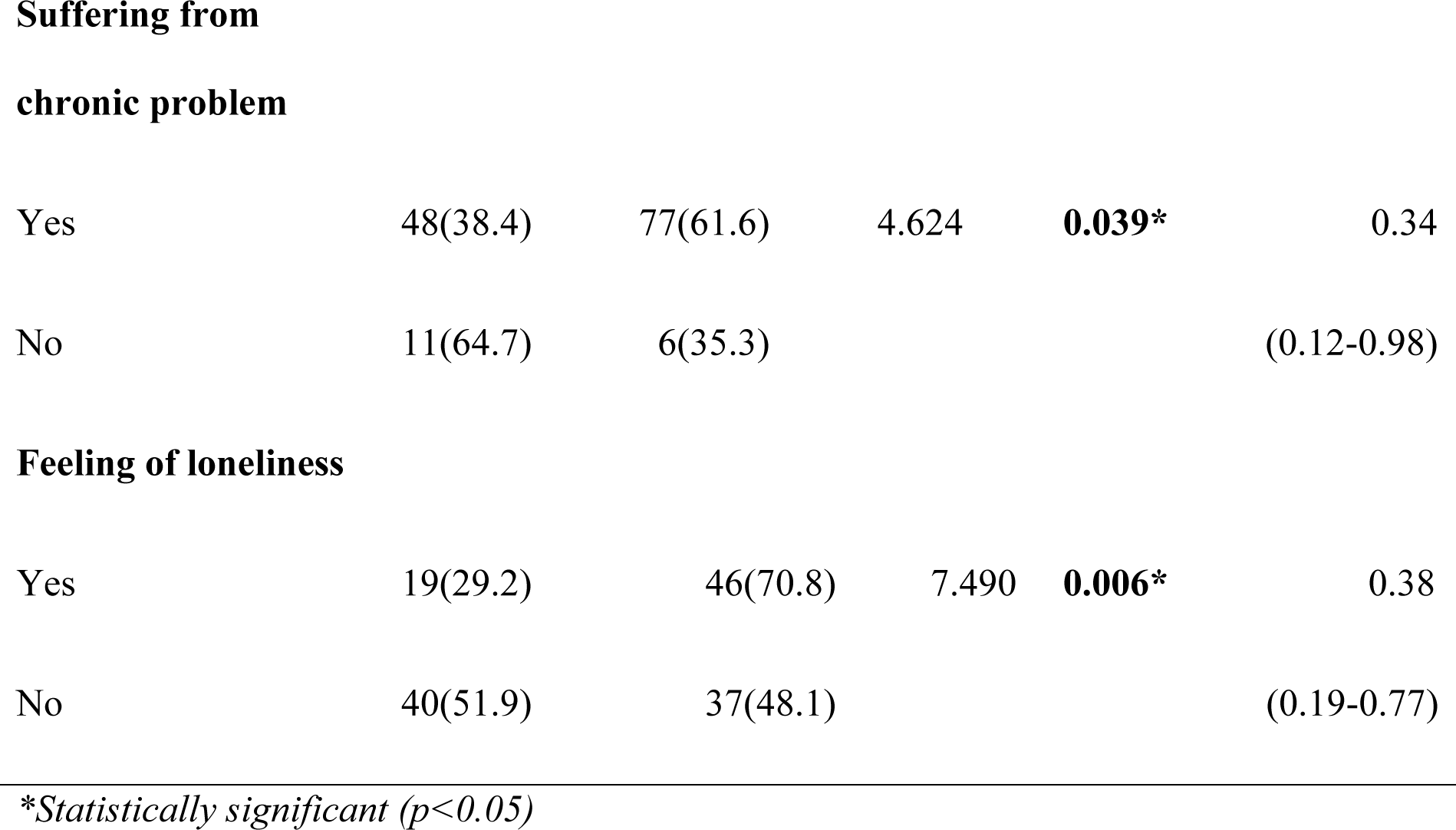
Association between depressive symptoms and risk factors n=142.

| Characteristics | Normal<br>n (%) | Depressive<br>Symptoms<br>n(%) | $\chi^2$<br>value | p<br>value | OR<br>(95% CI) |
| --- | --- | --- | --- | --- | --- |
| **Age** |  |  |  |  |  |
| 60-79 years | 44(48.4) | 47(51.6) | 4.828 | **0.028\*** | 2.25 |
| 80 and above | 15(29.4) | 36(70.6) |  |  | (1.08-4.66) |
| **Sex** |  |  |  |  |  |
| Male | 28(54.9) | 23(45.1) | 5.843 | **0.016\*** | 2.36 |
| Female | 31(34.1) | 60(65.9) |  |  | (1.17-4.75) |
| **Ethnicity** |  |  |  |  |  |
| Janajati | 32(47.8) | 35(52.2) | 2.016 | 0.156 | 1.63 |
| Brahmin Chhetri | 27(36) | 48(64) |  |  | (0.83-3.18) |
| **Marital status** |  |  |  |  |  |
| Single | 8(36.4) | 14(63.6) | 1.328 | 0.722 |  |
| Married | 7(41.2) | 10(58.8) |  |  |  |
| Widow/widower | 39(44.8) | 48(55.2) |  |  |  |
| Separated/divorced | 5(31.3) | 11(68.8) |  |  |  |
| **Education status** |  |  |  |  |  |
| Can read and write | 5(35.7) | 9(64.3) | 0.218 | 0.641 | 0.76 |
| Cannot read and write | 54(42.2) | 74(57.8) |  |  | (0.24-2.40) |
| **Past family type** |  |  |  |  |  |
| Nuclear | 19(30.6) | 43(69.4) | 5.388 | **0.020\*** | 0.44 |
| Joint | 40(50) | 40(50) |  |  | (0.22-0.89) |
| **Source of income** |  |  |  |  |  |
| Yes | 35(43.2) | 46(56.8) | 0.214 | 0.644 | 1.17 |
| No | 24(39.3) | 37(60.7) |  |  | (0.60-2.31) |
| **Suffering from chronic problem** |  |  |  |  |  |
| Yes | 48(38.4) | 77(61.6) | 4.624 | **0.039\*** | 0.34 |
| No | 11(64.7) | 6(35.3) |  |  | (0.12-0.98) |
| **Feeling of loneliness** |  |  |  |  |  |
| Yes | 19(29.2) | 46(70.8) | 7.490 | **0.006\*** | 0.38 |
| No | 40(51.9) | 37(48.1) |  |  | (0.19-0.77) |

*\*Statistically significant (p<0.05)*

Suffering from a chronic physical health problem (p=0.039) was found to be statistically significant for depressive symptoms. Elderly who suffered from chronic physical health problems were found to be more prone to have depressive symptoms (OR = 0.34, 95% CI: 0.12-0.98). The feeling of loneliness (p=0.006) was found to be statistically significant with depressive symptoms. Having feelings of loneliness made them vulnerable to depressive symptoms (OR = 0.38, 95% CI: 0.19-0.77s).

## Discussion

Depression is an important public health problem but the stigma and ignorance lingering around it prevents the true statistics of depression from being studied. Early identification and diagnosis of depressive symptoms help improve quality of life of the sufferer. Through this study, we aimed to determine the prevalence of depressive symptoms among the elderly living in old age homes of Kathmandu and ascertain the association between depressive symptoms and study characteristics. Our study shows association of depressive symptoms with age, sex, past family type, chronic physical health problems and feelings of loneliness.

58.5% of the respondents in our study were found to have depressive symptoms. Regarding its severity, mild depression was the most common (38.7%), followed by moderate symptoms (16.2%) while 3.5% of the elderly had severe symptoms. Another study conducted among the elderly in old age homes of Kathmandu valley by Kafle et al had found the prevalence of depression to be 47.3%, of which 34% had mild depression and 13.3% had severe depression(14). These variations could be due to variation in the study settings and coverage of a larger study area. Another study looking at the association of depression with the quality of life among the elderly had revealed the prevalence of depression to be 39.6%(15). Increased prevalence rate in our study could be a reflection of the impacts of COVID-19 pandemic which led to mobility restrictions imposed by the old age homes. This seems to be slightly higher than the rates of depression in India, Bangladesh(16,17).

Our study reports a higher prevalence of depressive symptoms among the females. This finding is consistent with other studies in this realm.(10,15). Increasing age was considered an important predictor of increasing depression in a study conducted by Zou et al among the inpatients of a tertiary care center in China(18). Being a female had greater association with depressive symptoms as suggested by other studies too(10,12,19). This might also be a result of lesser opportunities to females and them being more susceptible to discrimination in our society, which leads to a lower self-esteem and generates a feeling of helplessness.

Past family type was also identified as an important predictor of depressive symptoms. Only 50% of the participants who formerly were part of a joint family had depressive symptoms while 69.4% of those from nuclear families had depressive symptoms in our study. A possible reason behind this finding could be that those with joint families feel more satisfied and happy to have spent time with their family members in contrast to nuclear families. Similarly, there was a significant association between presence of chronic health problems and depressive symptoms. The association between chronic medical illnesses and depressive symptoms has also been established in other studies.(20,21). People with chronic ailments like arthritis, diabetes mellitus, heart diseases and chronic pulmonary diseases have been found to be at a higher risk of developing depression(21)The functional limitation brought about by these chronic illnesses are considered as the most important factor leading to depressive symptoms.(20).

There is also a close association between feelings of loneliness and depressive symptoms. Studies have shown that loneliness leads to serious health consequences; including depression. Those of higher age groups are reportedly found to have higher rates of loneliness due to loss of spouse, family, and social disengagement(22). This is largely due to having to live in old age homes after losing their family members, or partner or being separated from their family members. Loss of family members and abandonment ultimately render elderly with feelings of helplessness and loneliness.

Study characteristics like ethnicity, marital status, education and source of income were not found to have statistically significant association with depressive symptoms. In contrast, some studies have found ethnicity and education to be associated with depressive symptoms(19,23). A possible explanation for the variability is that these studies were community-based with participants from a diverse group of communities within the sample. There is limited literature exploring the association between marital status and source of income with depressive symptoms.

### Limitations

The results of our study should be interpreted in light of several limitations. As the study is conducted with a cross-sectional design, causal inference cannot be established. Face to face interview approach was adopted as a majority of the respondents did not know how to read and write. As the old age homes consist of a significant number of elderly living with disability, further research with appropriate methodology on them would be better. The outcomes of the study also cannot be generalized to any other region of the country.

### Conclusion

This study has revealed an alarming prevalence of depressive symptoms among the elderly living in old age homes of Kathmandu, which also be taken as a representation for the entire country. Timely identification and appropriately planned interventions can help reduce the burden. Policies, strategies and activities need to be introduced in collaboration with different health institutions to address the problem.

## Data availability

The datasets available and analyzed would be available from the corresponding author upon request.

## Acknowledgement

The authors would like to thank administrative offices, old age homes and all the participants of the study

